# Automated quantification of cerebral microbleeds for ARIA-H monitoring in Aging and Alzheimer’s Disease: A multicenter deep learning validation

**DOI:** 10.64898/2026.05.19.26353364

**Authors:** Zhen Xuen Brandon Low, Ella Rowsthorn, Mohamad-Reza Nazem-Zadeh, Michelle Francis, Catherine Robb, Robert Whiriskey, Maxwell Howcroft, Amy Brodtmann, John J. McNeil, Meng Law

**Author notes:** These authors contributed equally to this work (Zhen Xuen Brandon Low, Ella Rowsthorn).

## Abstract

We trained a self-configuring nnU-Net model for CMB segmentation in a heterogeneous multicenter sample (*n*=264), including 1.5T and 3T field strengths, SWI and T2*-GRE sequences, and community and clinical cohorts. Model performance was evaluated using 5-fold cross-validation with a focus on object-level detection metrics. Real-world performance was evaluated on scans from an unseen dataset of people with cerebrovascular disease (*n*=20). The model achieved 0.82 cluster Dice, 0.88 precision, and 0.77 sensitivity on hold-out test data. Notably, the model demonstrated a low false-positive rate, averaging 0.58 false positives (FPs) per scan, an improvement on existing publicly available models. The model achieved high performance in dataset of those with Alzheimer’s disease and mild cognitive impairment (0.89 cluster Dice, 0.94 sensitivity), supporting its utility in clinical settings where ARIA-H monitoring is critical. In external validation, the model maintained high robustness with 0.79 sensitivity and 0.95 FPs per scan. By leveraging a heterogenous training strategy and a self-adapting architecture, we demonstrate that deep learning can achieve high-precision CMB detection that is robust to domain shifts. The low FP rate suggests this publicly available pipeline is suitable for automated screening and lesion counting in heterogenous large-scale clinical trials, reducing the burden of manual quantification.

## 1 Introduction

Cerebral microbleeds (CMBs) are small, round, hypointense lesions ranging from 2 to 10 mm in diameter, visualized on T2*-weighted gradient-recalled echo (GRE) or susceptibility-weighted imaging (SWI) sequences [30]. Physiologically, they are formed following blood leakage from fragile small vessels resulting in perivascular hemosiderin-laden macrophages[8]. Underlying pathologies of CMBs include cerebral amyloid angiopathy (CAA) [30], monogenic small vessel disease (SVD) and hypertensive arteriopathy, and pose a risk of future intracerebral hemorrhage and siderosis [1, 3, 30].

The accurate detection and quantification of CMBs have moved beyond the diagnostic realm to treatment monitoring. The monitoring of Amyloid-Related Imaging Abnormalities (ARIA) is now a mandatory safety protocol [6] for people receiving amyloid lowering therapies with anti-amyloid monoclonal anti-bodies. ARIA can manifest as edema (ARIA-E) and hemorrhage (ARIA-H), including microhemorrhages. Consequently, the demand for precise longitudinal tracking of CMBs has surged. However, the current gold standard approach for quantification is manual identification by expert radiologists, imposing significant time burden and unnecessary use of expert resources. Furthermore, CMB identification is cognitively demanding and susceptible to radiologist fatigue [14, 23], with inter-rater reliability often yielding Kappa scores below 0.70 due to the difficulty of distinguishing indeterminate lesions from image noise [5]. The sheer volumes of 3D neuroimaging data in ‘big data’ cohorts often render manual segmentation unrealistic.

Automating CMB detection is difficult due to two key challenges: extreme class imbalance and radiological mimicry. In a typical MRI scan, CMBs occupy less than 0.01% of total brain volume, presenting an extreme class imbalance that biases standard deep learning loss functions toward predicting background class to minimize global error [7]. Furthermore, CMBs lack a unique intensity signature on susceptibility-sensitive sequences; healthy venous structures, basal ganglia calcifications, and cavernous malformations often exhibit hypo-intensities, indistinguishable from CMBs based on signal intensity alone. Distinguishing a spherical microbleed from the cross-section of a vein or a focal calcification requires a model capable of analyzing 3D volumetric context to discern linear continuity from spherical isolation [16]. Early approaches relied on two-stage pipelines, in which candidate proposal systems were followed by secondary false-positive reduction classifiers. While effective in principle, these methods are often computationally expensive and struggle to achieve an optimal balance between sensitivity and precision [4].

There has been a shift toward single-stage fully convolutional networks, such as the 3D U-Net, which allow for end-to-end segmentation [2]. However, a pervasive issue in prior studies is the lack of generalizability across domain shifts. Deep learning models trained on single-center data frequently fail when applied to external datasets due to variations in acquisition protocols (e.g., 1.5T vs. 3T field strength) and voxel anisotropy. Recent efforts to address this have attempted to improve robustness by training on multi-protocol data [25]. Yet, this broader applicability often comes at the cost of precision; state-of-the-art benchmarks in this domain report object-level F1 scores (Cluster Dice) of approximately 0.74, with sensitivities as low as 0.67 [25]. Similarly, while self-configuring networks have demonstrated efficacy on specific challenge datasets [15], they often lack validation on broad, heterogeneous cohorts that span the spectrum from healthy aging to advanced neurodegenerative pathology. We postulated that these performance limitations stem not from a lack of architectural complexity, but from the rigidity of static hyperparameters that fail to adapt to the varying resolutions and intensity distributions inherent in multicenter data.

We propose a highly robust, automated segmentation pipeline based on the self-configuring nnU-Net framework [13], validated on a heterogeneous multicenter cohort (*n*=264). Unlike previous studies restricted to single-domain data, our cohort integrates three distinct datasets: ASPREE (healthy elderly), VALDO (mixed vascular pathology), and AIBL (Alzheimer’s disease dementia and mild cognitive impairment (MCI)). This combination encompasses multi-domain heterogeneity, including variations in magnetic field strength (1.5T vs. 3T), sequence types (SWI vs. T2*-GRE) and clinical phenotype, effectively training the model to learn a unified representation of a ‘CMB’ despite significant variance in scanner hardware and image contrast. We hypothesized that such a model would perform well across datasets with heterogenous image acquisition protocols and clinical phenotypes, with especially high-performance in real-world testing relative to existing publicly available models.

## 2 Methods

### 2.1 Study Population and Data Acquisition

This retrospective multicenter study utilized a heterogeneous sample of 264 scans from three datasets (ASPREE, AIBL, VALDO) for training and 20 scans for external validation. All data were de-identified prior to analysis. Detailed acquisition parameters and demographic characteristics are summarized in Table 1.

**Table 1.**
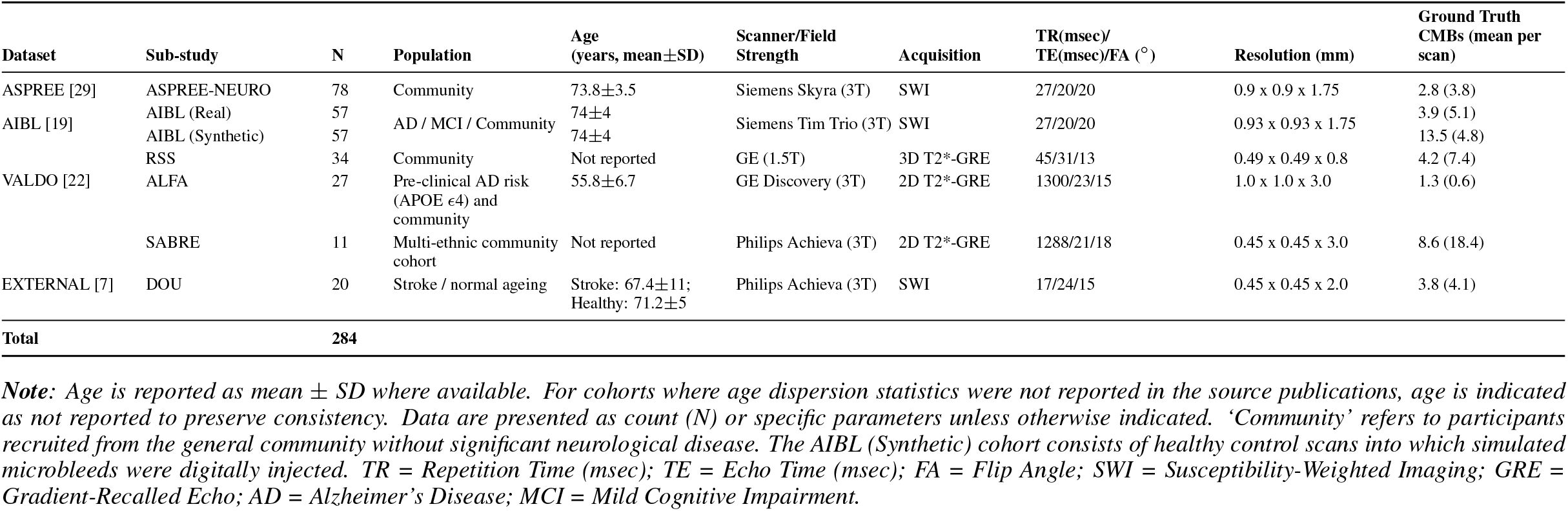
Demographic and imaging acquisition characteristics of the multicenter study cohort (n=284). The dataset comprises a heterogeneous mix of healthy aging, pre-clinical Alzheimer’s, and mixed vascular pathology participants whose scans were acquired across varying magnetic field strengths (1.5T and 3T) and scanner vendors (Siemens, GE, Philips). Note the significant domain shift introduced by the VALDO sub-studies, which utilize varying voxel geometries ranging from high-resolution 3D acquisitions to thick-slice 2D sequences.

#### ASPREE

We included 78 scans from 26 participants in the ASPREE-NEURO sub-study [29]. The study was approved by the Monash University Human Research Ethics Committee (Project Number: 2012001223), and all participants provided written informed consent. Scans were selected from three timepoints (baseline, 1-year, and 3-year follow-up) to introduce aleatoric variance (e.g. positioning) into the training distribution [27]. Data partitioning was participant-stratified to ensure all longitudinal timepoints from a given participant remained in the same fold. The ASPREE dataset has been separately and independently evaluated for CMBs by an experienced radiologist, and is a known ‘low count’ dataset.

#### AIBL

Data were obtained from the CSIRO Data Access Portal (Collection 50304) and is derived from the Australian Imaging, Biomarker & Lifestyle (AIBL) study, approved by the Austin Health and St Vincent’s Health Human Research Ethics Committees [19], with written informed consent. All available MRI scans within this collection with confirmed cerebral microbleeds (n = 57) were included; scan-level AD/MCI/community diagnostic labels were not available for this subset. To enhance training diversity, we included an additional 57 scans from the “rsCMB” dataset [19], containing synthetic microbleeds digitally injected via a physics-based simulation (total *n*=114). To prevent prevalence bias, we utilized the first version of the synthetic generation pipeline and performed a random selection of these participants to strictly match the sample size of the real pathology group. Because synthetic lesions may not fully capture the biological and imaging variability of endogenous CMBs, we additionally performed sensitivity analyses separating real and synthetic AIBL scans.

#### VALDO

We utilized the training set (*n*=72) from the MICCAI 2021 *Where is VALDO?* Challenge [22]. This dataset aggregates data from three epidemiological cohorts [12, 18, 24] (Table 1), comprising older adults from community-based ageing studies, including individuals without dementia and cognitively normal participants enriched for Alzheimer’s disease familial/genetic risk. This diversity provides high heterogeneity in voxel resolution (isotropic to 3.0 mm) and field strength. Inclusion of this data facilitates model robustness to scanner inter-variability. The overall study design, including the integration of these heterogeneous datasets through a unified preprocessing and deep learning pipeline, is presented in Figure 1.

**Figure 1:**
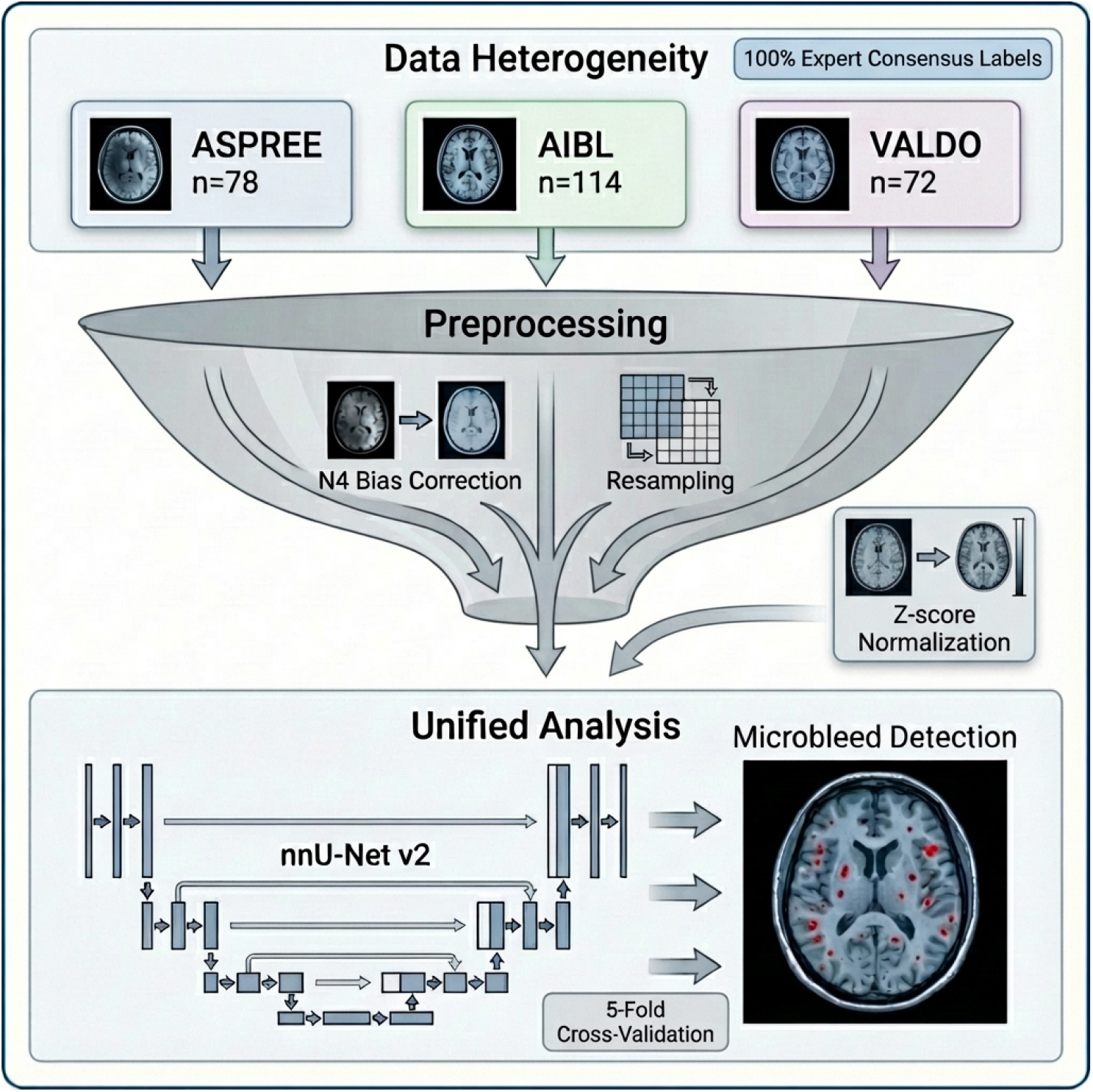
Multicenter Data Harmonization and Deep Learning Pipeline. (Top) Data Heterogeneity: The study integrates three distinct cohorts (ASPREE, AIBL, VALDO) totaling 284 scans (including external validation), representing heterogenous data including varying clinical phenotypes (Healthy Aging vs. Alzheimer’s disease dementia), magnetic field strengths (1.5T vs. 3T), and acquisition (T2*-GRE vs. SWI). All samples underwent a rigorous reference standard definition including 100% expert consensus labeling. (Middle) Unified Preprocessing: To address acquisition variance, a “funnel” approach harmonized the inputs via N4 bias field correction, resampling to a standardized 1.0mm isotropic resolution, and z-score normalization. (Bottom) Analysis Architecture: The harmonized volumes were fed into a self-configuring nnU-Net v2 architecture. Training utilized a stratified 5-fold cross-validation scheme on the internal cohort (n = 264) to ensure balanced exposure to all domains, resulting in voxel-wise microbleed detection (shown with representative prediction).

#### External Validation

To assess cross-site generalizability, we utilized an independent benchmark dataset (*n*=20) [7] comprising a balanced set of stroke patients and healthy aging controls (Table 1). This cohort introduces a domain shift in terms of scanner vendor (Philips) and voxel geometry (high in-plane resolution with anisotropic slice thickness) compared to the training data.

### 2.2 Reference Labels

Reference labels were standardized across datasets through a unified pipeline and annotated in ITK-SNAP (version 3.6.0) [31]. For ASPREE, two trained raters (MRNZ, ZXBL; >2 years’ experience) manually segmented CMBs using MARS criteria [9]. For VALDO, existing voxel annotations [22] underwent secondary quality control by ZXBL. For AIBL [19] and the external cohort [7], provided coordinate centroids were converted into volumetric masks by ZXBL, who manually delineated lesion boundaries. The final segmentations for all 284 scans were verified by two expert neuroradiologists (RW, MH; >12 years’ experience) to ensure a consistent clinical standard. Lesion counts were calculated from the final masks using 3D connected-component analysis (26-neighbor connectivity).

### 2.3 Data Harmonization and Preprocessing

A standardized preprocessing pipeline was applied to all scans. First, brain extraction was performed using FreeSurfer SynthStrip [11], and the resulting brain masks were dilated by 2 voxels. Bias field correction was applied using the N4ITK algorithm [26] via SimpleITK [17]. All scans were resampled to a standardized 1.0 mm isotropic resolution using third-order spline interpolation for images and nearest-neighbor interpolation for masks. Finally, intensities were clipped to the 0.5– percentile range and z-score normalized.

### 2.4 Model Architecture and Training

We employed the nnU-Net framework (Version 2, 3d_fullres configuration) [13] implemented in PyTorch [20]. Training utilized a dataset-stratified 5-fold cross-validation scheme on the development set (80% of total data) to maintain cohort balance. The model was optimized using stochastic gradient descent (initial learning rate of 1 *×* 10^*−*4^, poly decay) and a combined Dice and Cross-Entropy loss with deep supervision. Training proceeded for 1000 epochs on an NVIDIA A40 GPU using default on-the-fly data augmentation. Inference utilized a sliding-window approach with Gaussian weighting and no minimum cluster size filtering. For the external validation set, predictions were generated using an ensemble of the five cross-validation models. Automated lesion counts were calculated via 3D connected component analysis (26-neighbor connectivity). For evaluation, a true positive was defined as any predicted cluster overlapping a ground truth lesion by at least one voxel.

### 2.5 Inference and Statistical Analysis

Performance was evaluated on the held-out test sets from the stratified 5-fold cross-validation. Metrics included lesion-level Sensitivity (Recall), Precision (Positive Predictive Value), mean False Positives (FPs) per scan, and Cluster Dice. In addition, a scan-level burden classification analysis was performed using a clinically relevant threshold (≤ 4 vs >4 CMBs) [6, 21], and corresponding positive and negative predictive values (PPV, NPV) were calculated. For negative controls (scans with zero ground truth lesions), correct rejection was assigned to a Sensitivity and Precision of 1.0. Due to non-normal distributions driven by lesion sparsity, results are reported as both mean (standard deviation) and median [interquartile range], with 95% confidence intervals generated via subject-level bootstrapping (1,000 resamples). A sensitivity analysis was conducted to compare performance between endogenous (“real”) and synthetic lesions in the AIBL cohort. Finally, agreement between manual and automated clinical lesion counts was assessed using Spearman’s rank correlation (*ρ*). Statistical analyses were performed using Python 3.9 with the scipy [28] and numpy [10] libraries.

### 2.6 Code Availability

The MedNet-CMB implementation is publicly available at https://github.com/iBrain-Lab/MedNet-CMB.

## 3 Results

### 3.1 Model Performance - Quantitative

Across all scans (*n*=264), the model achieved an object-level Precision of 0.88 and a Sensitivity of 0.77, resulting in an F1-score of 0.82 (Table 2). The model produced an average of 0.58 False Positives (FPs) per scan (Figure 2), indicating a relatively low false-positive burden.

**Table 2.**
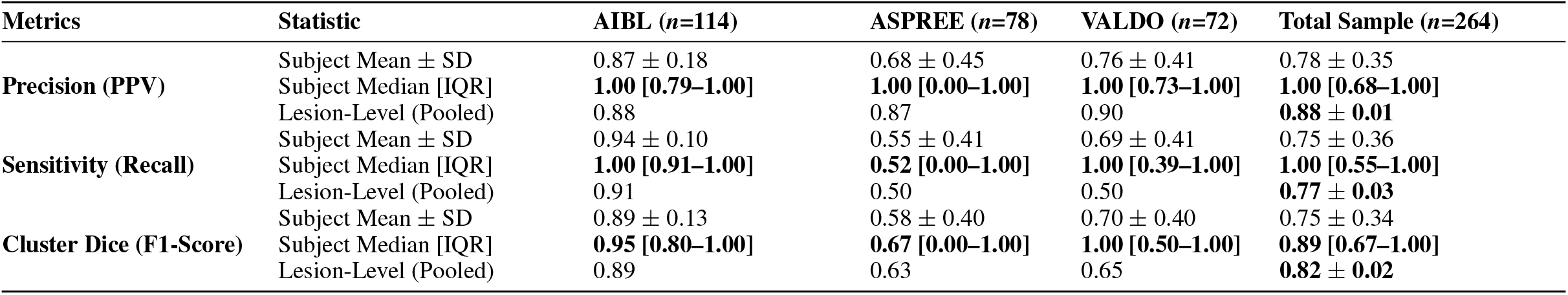
Detection Performance. Metric-based comparison across cohorts. Data are reported at the Subject-Level (Macro-average: Mean ± SD and Median [IQR]) to demonstrate per-patient reliability. Lesion-Level (Pooled) metrics were calculated across all lesions. Pooled SDs for lesion-level metrics in the Total Sample were estimated via bootstrapping (n=1,000).

| Metrics | Statistic | AIBL ($n=114$) | ASPRE (n=78) | VALDO ($n=72$) | Total Sample ($n=264$) |
| --- | --- | --- | --- | --- | --- |
| **Precision (PPV)** | Subject Mean ± SD | 0.87 ± 0.18 | 0.68 ± 0.45 | 0.76 ± 0.41 | 0.78 ± 0.35 |
|  | Subject Median [IQR] | **1.00 [0.79–1.00]** | **1.00 [0.00–1.00]** | **1.00 [0.73–1.00]** | **1.00 [0.68–1.00]** |
|  | Lesion-Level (Pooled) | 0.88 | 0.87 | 0.90 | **0.88 ± 0.01** |
| **Sensitivity (Recall)** | Subject Mean ± SD | 0.94 ± 0.10 | 0.55 ± 0.41 | 0.69 ± 0.41 | 0.75 ± 0.36 |
|  | Subject Median [IQR] | **1.00 [0.91–1.00]** | **0.52 [0.00–1.00]** | **1.00 [0.39–1.00]** | **1.00 [0.55–1.00]** |
|  | Lesion-Level (Pooled) | 0.91 | 0.50 | 0.50 | **0.77 ± 0.03** |
| **Cluster Dice (F1-Score)** | Subject Mean ± SD | 0.89 ± 0.13 | 0.58 ± 0.40 | 0.70 ± 0.40 | 0.75 ± 0.34 |
|  | Subject Median [IQR] | **0.95 [0.80–1.00]** | **0.67 [0.00–1.00]** | **1.00 [0.50–1.00]** | **0.89 [0.67–1.00]** |
|  | Lesion-Level (Pooled) | 0.89 | 0.63 | 0.65 | **0.82 ± 0.02** |

**Figure 2:**
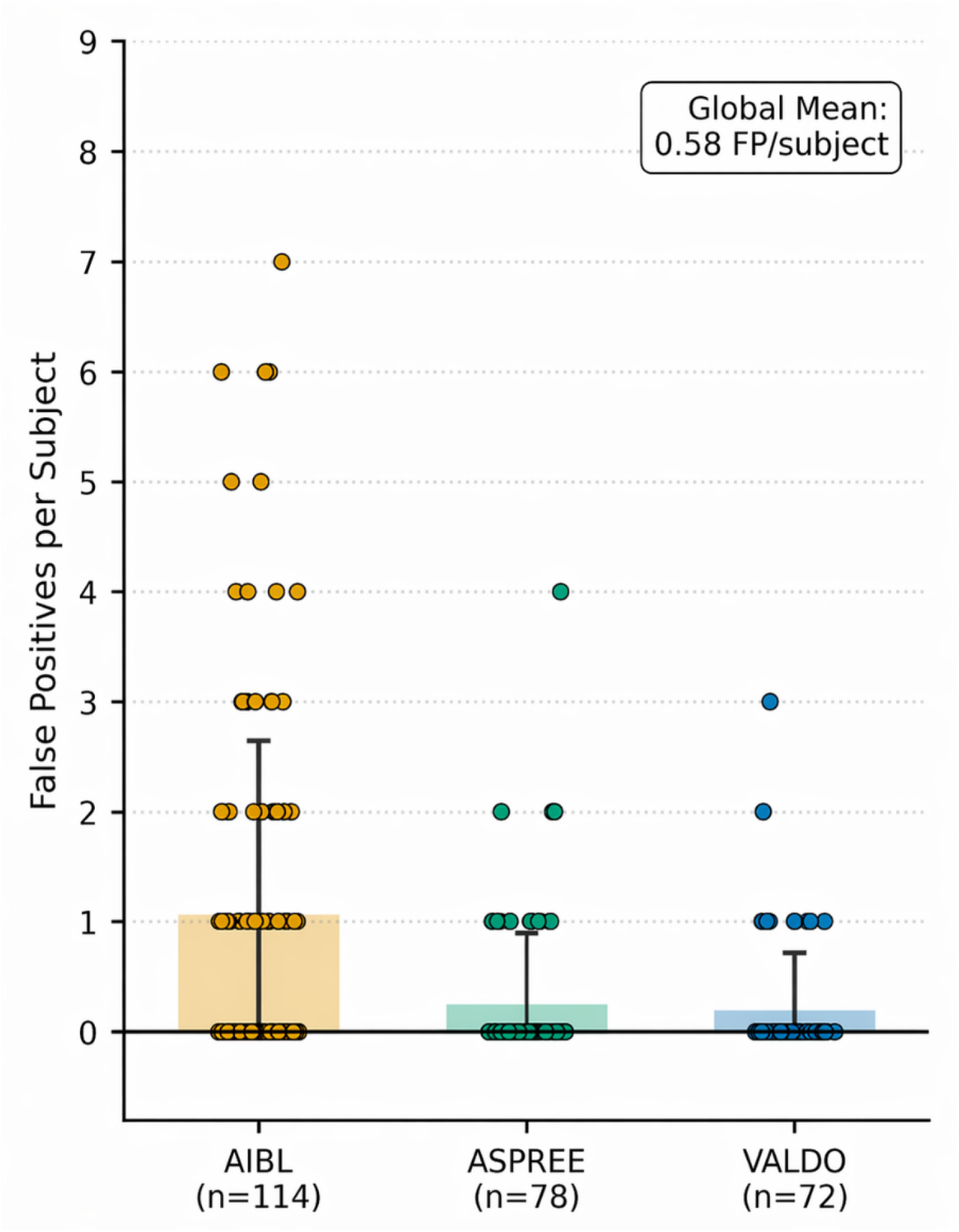
Distribution of False Positives (FPs) per scan across test cohorts. The strip plot visualizes individual scan FP counts (jittered points) overlaid with bars representing the mean FP rate for each dataset. Error bars indicate the standard deviation. Despite variations in acquisition protocols, the model maintains high specificity with a global mean of 0.58 FPs per scan, with the majority of scans in all cohorts yielding zero false positives.

Subgroup analysis revealed the highest performance in the AIBL cohort (*n*=114, AD/MCI population), with a Sensitivity of 0.94 and a Precision of 0.87. In the ASPREE cohort (*n*=78), Precision was 0.68, though Sensitivity was lower at 0.55. The heterogeneous VALDO cohort (*n*=72) had a Sensitivity of 0.69 and a Precision of 0.76. Post-hoc stratification of the VALDO cohort indicated that voxel geometry was a critical performance determinant: the ALFA (3T, 3.0 mm slices) and RSS (3T, 0.8 mm slices) sub-cohorts achieved sensitivities of 0.70 and 0.71 respectively, whereas the SABRE sub-cohort (3T), characterized by extreme voxel anisotropy (0.45 *×* 0.45 mm in-plane vs. 3.0 mm through-plane), yielded the lowest sensitivity (0.56). As an exploratory scan-level burden analysis, classification at the clinically relevant threshold of ≤ 4 versus >4 CMBs achieved strong performance in the internal cohort (PPV=0.96, NPV=0.93), with robust dataset-specific performance in AIBL (PPV=0.95, NPV=0.98), ASPREE (PPV=1.00, NPV=0.89), and VALDO (PPV=1.00, NPV=0.95).

### 3.2 Model Performance - Qualitative

Qualitative inspection (Figure 3) demonstrated high fidelity in segmenting lobar microbleeds, achieving near-perfect pixel-level overlap with ground truth (Figure 3A). Notably, robustness was observed even in the posterior fossa (Figure 3B), a region frequently obscured by skull-base artifacts. In several instances, the model detected deep, focal hypo-intensities absent in the initial annotations that were retrospectively adjudicated as true positives (“AI-assisted discovery,” Figure 3C). However, the model occasionally failed to segment faint satellite lesions adjacent to dominant microbleeds (Figure 3D).

**Figure 3:**
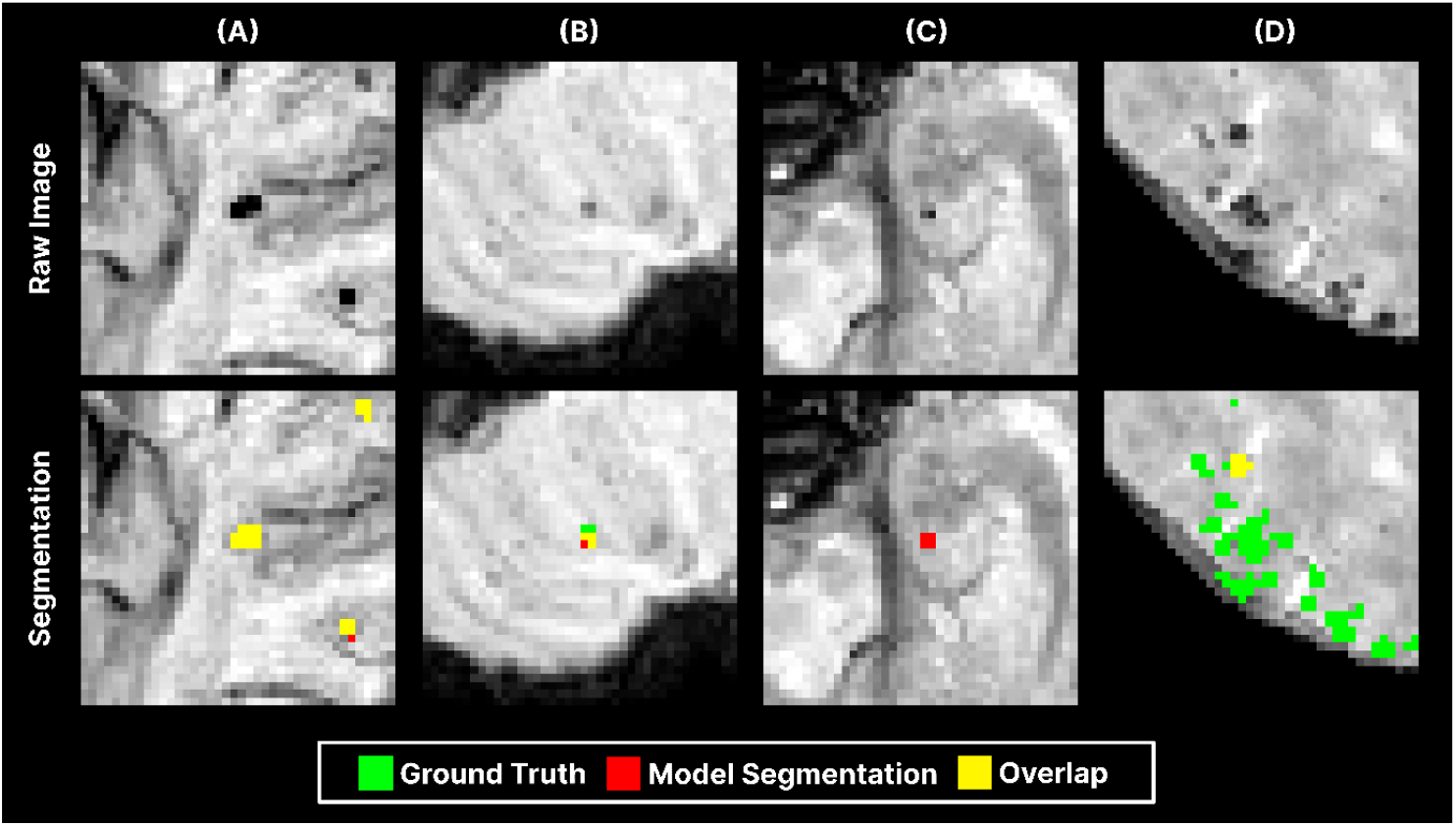
Qualitative assessment of segmentation performance. The top row displays the raw Minimum Intensity Projection (MinIP) MRI crops, and the bottom row displays the corresponding segmentation overlays where green indicates the human ground truth, red denotes the model prediction, and yellow represents the overlap (agreement). (A) High Fidelity: The model demonstrates precise pixel-level agreement with expert annotation on a clear lobar microbleed. (B) Deep Structure Robustness: The model successfully detects a cerebellar lesion (yellow) despite fragmentation, correctly localizing pathology in a region prone to susceptibility artifacts. (C) AI-Assisted Discovery: A deep focal lesion is detected by the model (red) but was absent in the initial annotation. Retrospective expert review confirmed this as a true positive, highlighting the utility of the model in drawing attention to subtle pathology. (D) Sensitivity Threshold: In a cluster of lesions, the model successfully identifies the dominant microbleed (yellow) but fails to segment the fainter satellite lesions (green), illustrating a contrast-dependent sensitivity threshold.

### 3.3 Model Performance – Real-world External Validation

On the independent external validation dataset (*n*=20), the model achieved a mean object-level Precision of 0.83 and a Sensitivity of 0.79 (F1-score: 0.76). The false positive rate was 0.95 FPs/scan, maintaining sub-one false positive performance despite the domain shift in scanner vendor and acquisition protocol. At the same scan-level threshold, external validation yielded a PPV of 1.00 and NPV of 1.00.

### 3.4 Sensitivity Analysis: Real vs. Synthetic Scans

Performance on synthetic AIBL data (Mean F1: 0.92) was comparable to endogenous “real” pathology (Mean F1: 0.86). This consistency suggests the model utilized morphological features common to both real and simulated lesions rather than overfitting to synthetic artifacts. When synthetic AIBL scans were excluded, aggregate performance decreased (Precision=0.83, Sensitivity=0.62, Mean F1=0.71), but remained within the range reported in prior studies.

### 3.5 Clinical Burden Quantification

Automated lesion counts demonstrated strong rank-order correlation with manual counts across the full cohort (Spearman *ρ*=0.85, p<0.001; Figure 4). When stratified by dataset, concordance was highest in the AIBL cohort (*ρ*=0.94) and robust in the healthy ASPREE cohort (*ρ*=0.57). Correlation was moderate in the VALDO dataset (*ρ*=0.73), where thick-slice acquisitions resulted in the underestimation of lesion counts in high-burden cases.

**Figure 4:**
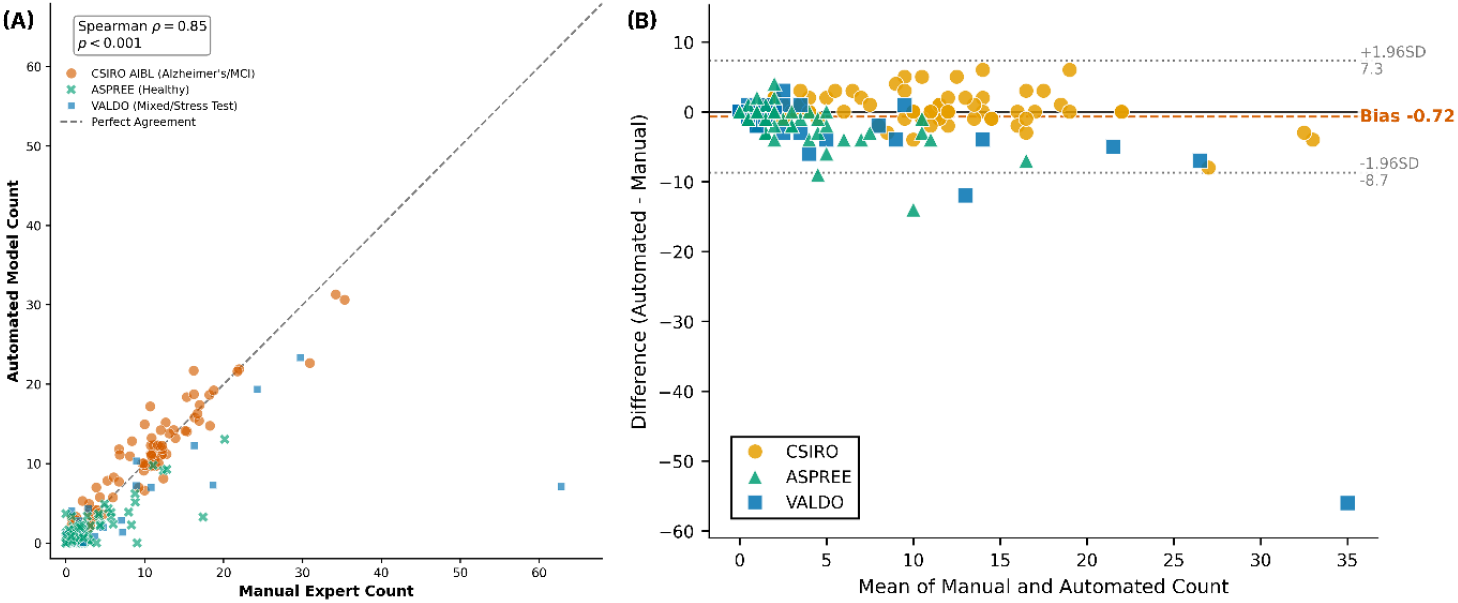
Agreement and bias analysis of automated microbleed counting. (A) Concordance Scatter Plot: Comparison of automated deep learning predictions vs. manual expert ground truth across the full cohort (n=264; Spearman *ρ* = 0.85, p<0.001). Points are jittered slightly to visualize density. The dashed diagonal line represents perfect agreement (y=x). The model demonstrates high reliability in the primary target population (AIBL, orange circles) and correctly classifies the majority of healthy controls (ASPREE, green markers) as having zero lesions. (B) Bland-Altman Plot: Assessment of measurement error showing the difference between methods (Automated - Manual) against the mean count. The orange dashed line indicates a minimal systematic bias of -0.72 lesions, while grey dotted lines represent the 95% limits of agreement.

## 4 Discussion

In this multicenter study, we trained a self-configuring deep learning framework to detect CMBs across a heterogeneous sample of MRI field strengths, acquisition parameters and clinical phenotypes. The central finding is the model’s excellent performance in a group of people with MCI and AD (AIBL), where it achieved a Sensitivity of 0.94 and a Precision of 0.87, with strong correlation to manual expert counts (*ρ*=0.94). Crucially, the model maintained a low false-positive rate (0.58 per scan) across diverse acquisition protocols, potentially mitigating the “alarm fatigue” that has historically hindered the adoption of automated screening tools in clinical practice.

The clinical importance for automated CMB quantification has increased with the implementation of anti-amyloid therapies, such as lecanemab and donanemab. The safety protocols for these immunotherapies mandate rigorous monitoring for ARIA-E and ARIA-H. In this context, precision is as critical as sensitivity; a false-positive detection could erroneously exclude a patient from therapies or classify someone as having excessive CMBs, placing them in the category of ARIA-H and potentially leading to the unnecessary suspension of a disease-modifying therapy. Our model’s high precision in the MCI/AD cohort, combined with its ability to correctly classify anatomical mimics, such as venous cross-sections and calcifications, suggest it is well-suited for this safety application.

Furthermore, the high concordance with expert counts implies that the model can reliably track longitudinal changes in lesion burden, alleviating a radiological bottleneck in both clinical trials and practice.

Our approach compares favorably with recent benchmarks. Early deep learning methods often suffered from prohibitive false-positive rates when applied to external data. Even recent multi-source frameworks, such as SHIVA-CMB [25], have reported sensitivities of approximately 0.67 on held-out test sets. In contrast, our pipeline achieved a higher aggregate sensitivity (0.77) and precision (0.88), with robust performance on the completely independent external validation set (Precision 0.83). This suggests that the quality of ground-truth annotation (specifically, rigorous human-in-the-loop verification) remains a decisive factor in model performance, potentially outweighing the sheer volume of training data.

A unique contribution of this study is the granular failure analysis within the heterogeneous VALDO cohort. Contrary to the assumption that field strength is the sole determinant of CMB detectability, our results highlight voxel isotropy as a governing factor for 3D deep learning performance. While 3T imaging generally outperformed 1.5T imaging, this advantage was nullified by extreme voxel anisotropy. The performance degradation in the SABRE sub-cohort (3T, but 3.0 mm slice thickness) demonstrates that the partial volume effects in thick slice 2D acquisitions obscure the spherical morphology required by 3D convolutional kernels to distinguish lesions from noise. This has practical implications for clinical trial design: to maximize the utility of AI-based monitoring, acquisition protocols should prioritize isotropic 3D sequences (like SWI) over field strength alone.

From a methodological perspective, these results validate the efficacy of data-driven architectural configuration over static model design. Previous deep learning approaches for CMB detection have typically relied on fixed hyperparameter sets (e.g., fixed patch sizes and pooling kernels) optimized for a specific resolution. Our use of the self-configuring nnU-Net framework allowed the pipeline to dynamically adapt its topology, specifically the resampling strategy and receptive field, to the median characteristics of the aggregated training data. This adaptability likely contributed to the model’s ability to generalize across the extreme resolution disparities of the VALDO cohort without the need for manual architectural tuning.

A potential concern in deep learning studies utilizing synthetic data is the risk of the network overfitting to artificial fiducials, thereby failing to generalize to real biological targets. In our study, the high concordance between the ‘Real’ and ‘Synthetic’ AIBL sub-cohorts (Dice 0.86 vs. 0.92) mitigates this concern. We attribute this robustness to the underlying fidelity of the rsCMB simulation pipeline [19]. Unlike simplistic data augmentation techniques that merely insert hypointense circles, the rsCMB method utilizes a physics-based model to simulate the local magnetic susceptibility dipole and the resulting ‘blooming effect’ characteristic of paramagnetic hemosiderin. By training on these biophysically accurate representations, the network was forced to learn the complex volumetric signature of microbleeds, including phase shifts and signal loss patterns, rather than simple geometric shape descriptors. This ensures that the high sensitivity reported here reflects genuine proficiency in detecting cerebral pathology, rather than an artifact of the synthetic generation process.

The primary strength of this study is the rigorous construction of the training and validation cohorts, which encompassed the full spectrum of CMB presentations, from sparse lesions in healthy aging (ASPREE) to greater numbers in people with MCI/AD (AIBL) and difficult mimics in mixed-pathology datasets (VALDO). Additionally, the use of synthetic pathology (“rsCMB”) provided a controlled validation environment, confirming that the model learns true lesion features rather than dataset-specific artifacts. However, several limitations must be acknowledged. First, the study is retrospective; prospective validation in an active clinical trial workflow is required to further assess real-world utility. Second, performance on thick-slice 2D GRE scans remains suboptimal. While this reflects a physical limitation of the imaging modality, future work could explore super-resolution techniques or 2.5D architectures to improve detection in legacy datasets. Third, while we utilized expert radiological consensus as the ground truth, the study lacked pathological confirmation, which remains the absolute gold standard for distinguishing microbleeds from mimics. Finally, although our dataset was multicenter, it was heavily weighted towards research-quality scans; performance on “noisy” routine clinical scans requires further evaluation.

## 5 Conclusion

In this work, we validated a self-configuring deep learning framework for the automated quantification of cerebral microbleeds in a large, heterogeneous multicenter cohort. By leveraging a diverse training strategy that spans healthy aging, vascular pathology, and neurodegenerative disease, we demonstrated that the model achieves high-precision detection (PPV=0.88) robust to significant variations in scanner hardware and acquisition protocols.

Crucially, the model’s performance peaked in the MCI/AD cohort, achieving strong correlation with expert counts (*ρ*=0.94) and a minimal false-positive rate (0.58 per scan) across all datasets. These findings indicate that the pipeline effectively overcomes the specificity limitations of previous methods, support automated CMB screening, lesion counting, and triage of scans for expert radiological review in the era of amyloid-modifying immunotherapies. Future work will focus on integrating this tool into longitudinal clinical trial workflows to prospectively validate its utility in detecting incident ARIA-H.

## Data Availability

The AIBL dataset is publicly available via the CSIRO Data Access Portal (Collection 50304). The VALDO dataset is publicly available via the MICCAI 2021 Where is VALDO? Challenge. The external validation dataset is publicly available from Dou et al. (2016). Model weights and inference code are publicly available on GitHub at https://github.com/iBrain-Lab/MedNet-CMB.

